# “Understanding Clinical Variances in Urinary Isolates: Pathogenic vs. Non-Pathogenic Impacts in Tertiary Healthcare of North India”

**DOI:** 10.1101/2024.01.11.24301147

**Authors:** Bimalesh Yadav, Prasan Kumar Panda, Ravi Kant, Balram Ji Omar, Sandeep Saini, Vikas Kumar Panwar, Yogesh Arvind Bahurupi

## Abstract

**INTRODUCTION:** The clinical decision-making landscape in urinary tract infections (UTIs) hinges on discerning between pathogenic and non-pathogenic organisms. The accurate interpretation of culture results relies significantly on precise collection methods, the patient’s clinical status, and organism characteristics. This pioneering study, the first of its kind, systematically examines the clinical outcomes associated with pathogenic and non-pathogenic urinary isolates. Nestled within the tertiary healthcare context, this research not only fills a critical knowledge gap but also endeavors to unravel the economic and clinical implications that underscore the distinctive nature of UTIs in North India.

**OBJECTIVE:** To conduct a comprehensive assessment of the clinical and economic impacts of urinary tract infections (UTIs) caused by pathogenic and non-pathogenic organisms in North India’s tertiary healthcare. Evaluate hospitalization duration, antimicrobial usage, costs, mortality rates, and patient demographics to inform customized management strategies, optimize resource allocation, and enhance overall UTI patient care.

**METHODOLOGY:** Conducted at AIIMS Rishikesh, a longitudinal-exploratory study over 18 months included 275 participants meeting specific criteria. Data entry via Redcap ensured precision. SPSS (Version 23) facilitated statistical analysis encompassing chi-square tests and logistic regression, prioritizing ethical considerations and patient consent.

**RESULTS:** Pathogenic Cases: Comprising 90.54% of the cohort, average hospital stays of 14.68 days, and expenses of $29.12 on antibiotics with a mortality rate of 3.6%.

Pathogenic-Commensal Cases: Constituting 61.81% with an average stay of 13.88 days, antibiotic expenses at $28.46±9.33, and a mortality rate of 3.2%.

Pathogenic Colonizer Cases: Accounting for 14.18%, with an average stay of 14.67 days, antibiotic expenses at $27.69±11.02, and a mortality rate of 0.36%.

Direct Pathogenic Cases: Representing 14.54% with longer stays (18.08 days) and higher antibiotic expenses $33.28±5.01

Non-Pathogenic Cases: Makeup 9.45% with variable stays and expenses.

**DISCUSSION:** This ground-breaking study explores clinical variances in urinary isolates, distinguishing pathogenic (90.54%) and non-pathogenic cases (9.45%). Subtypes within pathogenic cases offer nuanced insights, guiding tailored patient care and optimizing resources. Non-pathogenic cases, though fewer, reveal variability, emphasizing the study’s depth. Recognizing the clinical impact of seemingly benign isolates becomes crucial amid rising antimicrobial resistance, urging judicious antimicrobial prescription.

**CONCLUSION:** This study pioneers insights into North India’s urinary isolates, emphasizing customized management. Distinguishing pathogenic from non-pathogenic cases, especially subtypes, is imperative. Identifying and avoiding unnecessary antimicrobial use emerge as pivotal interventions, contributing significantly to global antimicrobial resistance efforts and alleviating economic burdens on healthcare systems worldwide.

## INTRODUCTION

Within the intricate domain of infectious diseases, the accurate identification of microorganisms isolated in culture poses a formidable challenge for clinicians and microbiologists. The distinction between true pathogens necessitating immediate treatment, commensal organisms coexisting harmoniously within the host, colonizers in potential dormancy, and contaminants introduced during specimen collection is crucial. This study stands as the first of its kind, undertaking a comprehensive exploration into the clinical variances of urinary isolates. It uniquely compares the distinctions between pathogenic and non-pathogenic organisms, aiming to uncover economic and clinical implications within the tertiary healthcare landscape of North India.

Infection initiation occurs when disease-causing microbes enter the body and multiply, leading to cell damage and illness. A pathogen, fulfilling Koch’s postulates, is the definitive causative agent demanding prompt intervention. Conversely, non-pathogens do not warrant antimicrobial treatment, highlighting the need for precise identification. Commensal organisms engage in a symbiotic relationship without causing harm, while colonizers can transition into pathogens under specific conditions. Contaminants, introduced unintentionally during sample collection, pose diagnostic challenges [1].

Urine samples, crucial for clinical analysis, face contamination risks due to the diverse microorganisms in the urethra. Proper collection methods, such as voided midstream or catheterized specimens, are imperative to ensure reliable culture results. This study focuses on urinary tract infections (UTIs), emphasizing meticulous specimen collection and transportation for accurate diagnosis.

Microscopy plays a pivotal role, in detecting polymorphonuclear leukocytes in urine, indicating infection even without bacterial growth. The Kass Criteria set a bacterial count threshold for significant bacteriuria. Understanding the complexities of microbial identification becomes paramount, considering the challenges posed by factors like diagnostic uncertainty and patient demand.

The misuse of antimicrobials, especially in the context of viral infections, contributes significantly to antimicrobial resistance. Broad-spectrum antimicrobials, when unnecessary, further escalate resistance risks. This study addresses these challenges by proposing a stepwise exploratory model to distinguish pathogenic from non-pathogenic urinary isolates. By preventing unnecessary antimicrobial usage, the study aims to curb resistance and reduce the economic burden on healthcare systems.

The literature review underscores the global impact of UTIs, particularly catheter-associated urinary tract infections (CAUTIs), emphasizing their prevalence in healthcare settings. This study, a pioneering initiative, aims to contribute to the understanding of microbial complexities in UTIs. The stepwise exploratory model holds the potential to guide treatment decisions, optimize resources, and improve patient outcomes, aligning with the imperative to combat antimicrobial misuse and reduce the economic strain on the global healthcare landscape.

## METHODOLOGY

Following the approval from the Institutional Ethics Committee, AIIMS (DHR REG NO: EC/NEW/INST/UA/0180) and with approval number 295/IEC/PGM/2022, this study is conducted within the esteemed halls of AIIMS Rishikesh. The research adopts a prospective longitudinal-exploratory design, spanning 18 months and commencing in January 2022. The nucleus of this research comprises a meticulously selected sample of 275 participants meeting specific inclusion criteria. Data entry, facilitated through Redcap, is employed to ensure precision and reliability. The sophisticated SPSS software (Version 23) is utilized for statistical analysis, which includes chi-square tests and logistic regression. A paramount focus of our methodology is the assurance of confidentiality, coupled with a strong commitment to ethical considerations, notably prioritizing patient consent. This refined approach heralds a profound exploration into the intricacies of urinary tract infections within a hospital milieu, aiming to contribute valuable insights to the field.

**FIG 1.**
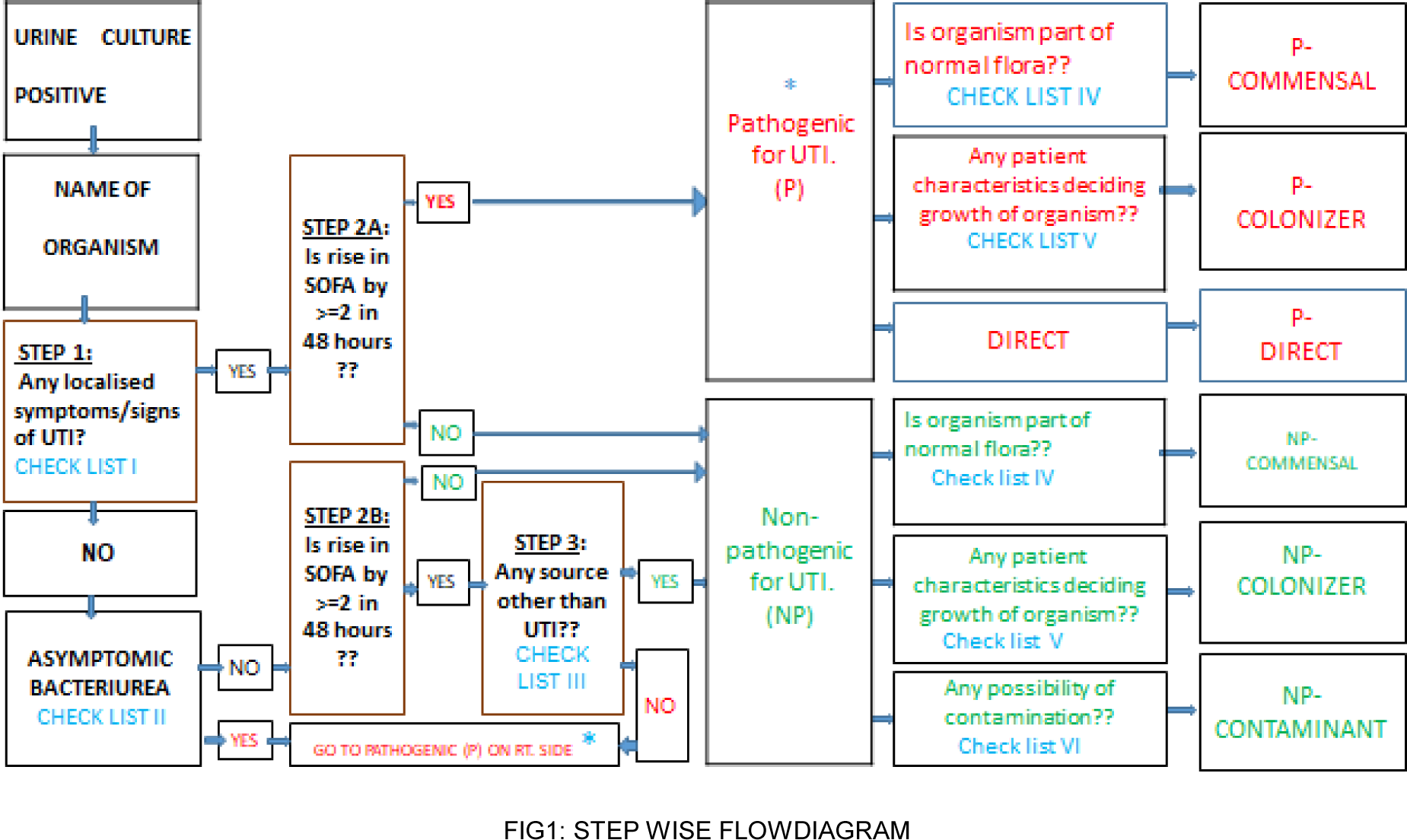
: **STEP WISE FLOWDIAGRAM**

## RESULT

The total cohort consisted of 275 individuals, and within this group, the average age of the cohort is reported at 47.66 ± 16.67 years, with a distinct gender distribution of 172 males to 103 females, constituting 62.5% and 37.5%, respectively. Noteworthy averages include a hospital stay duration of 13.93 days, an antibiotic (ABX) usage period of 6.75 days and an average expense on ABX of $27.48±10.68.

Within this cohort, a subset of cases is identified as pathogenic (N=249), representing a substantial 90.54%. This subgroup exhibits an average age of 47.32 ± 16.96 years, with a male-to-female ratio of 156 (62.7%) to 93 (37.3%). Key metrics for this group include an average duration of hospital stay at 14.68 ± 11.55 days, ABX usage of 6.81 ± 2.02 days, and an average expense on ABX amounting to $29.12± 9.22. The mortality rate within this pathogenic subgroup is reported as 3.6%, accounting for 10 cases.

The assessment of pathogenicity takes into account different perspectives: all cases are identified as pathogenic by microbiologists (100%), treating physicians categorize 272 cases (98.9%) as pathogenic, and an algorithm designates 249 cases (90.5%) as pathogenic.

Further categorization within the pathogenic cases reveals three subgroups: Commensal, Colonizer, and Direct. Commensal cases, constituting 61.81% of the pathogenic group, exhibit unique features, including an average expense on ABX of $28.46±9.33. Colonizer cases (14.18%) and Direct cases (14.54%) demonstrate distinct patterns in demographics, length of stay, ABX usage, and financial implications.

In contrast, the non-pathogenic cases (N=26) form 9.45% of the total cohort. This subgroup is characterized by an average age of 50.88 ± 13.42 years, with a male-to-female ratio of 16 (61.5%) to 10 (38.5%). The average length of hospital stay is 6.77 ± 3.06 days, the ABX usage duration is 6.23 ± 1.34 days, and the average expense on ABX is $11.79± 11.02. Notably, no mortality cases are recorded in this non-pathogenic group.

A more detailed breakdown of non-pathogenic cases into Commensal, Colonizer, and Contaminants reveals specific characteristics. Commensal cases, representing 6.9% of non-pathogenic instances, showcase an average expense on ABX of $11.59±11.07. Colonizer cases (1.81%) and Contaminants (0.72%) present their patterns in demographics, ABX usage, and financial implications.

**Figure 2:**
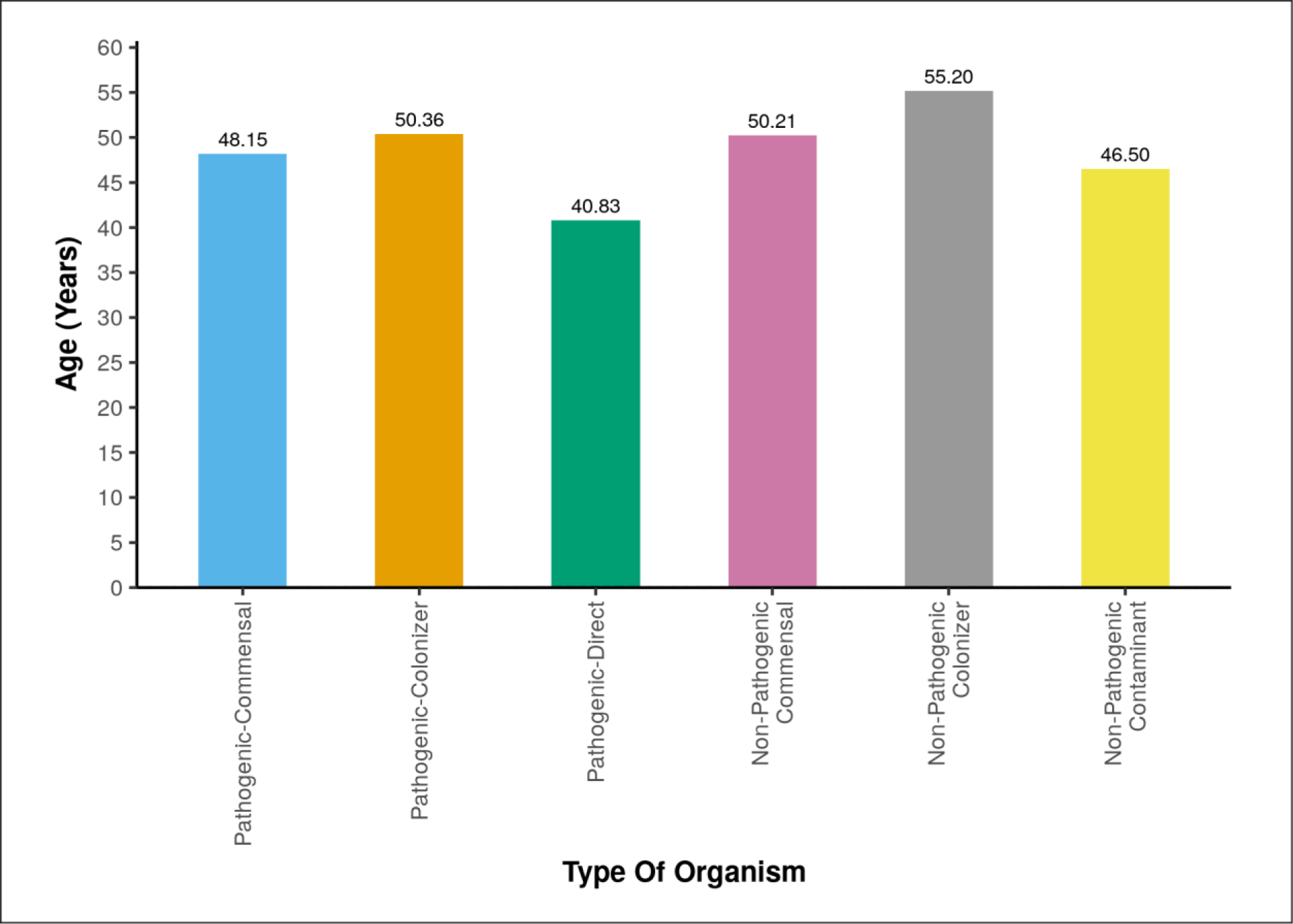
**Association Between Nature of Organism and age**

**Figure 3:**
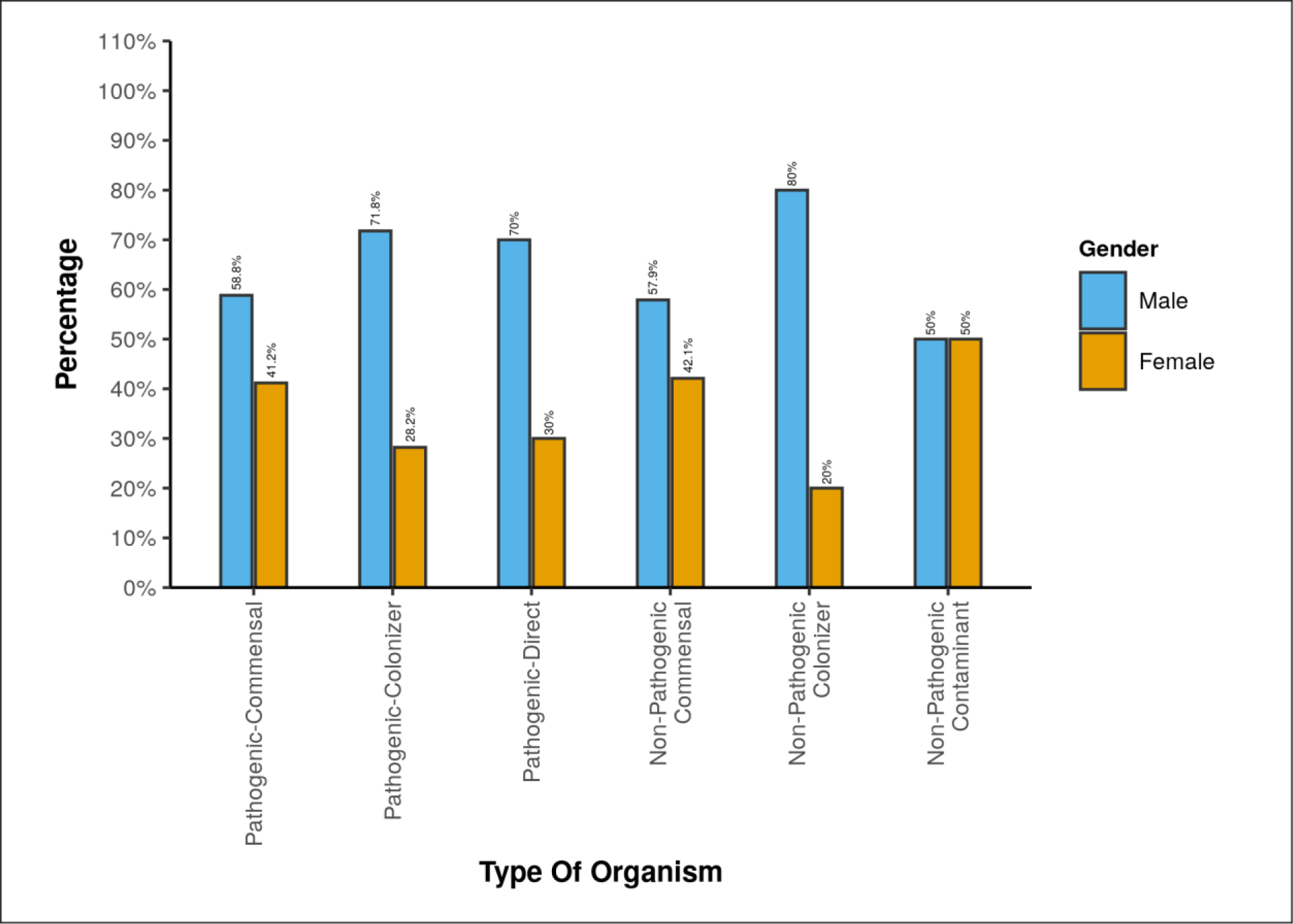
**Association Between Nature of Organism and gender**

**Figure 4:**
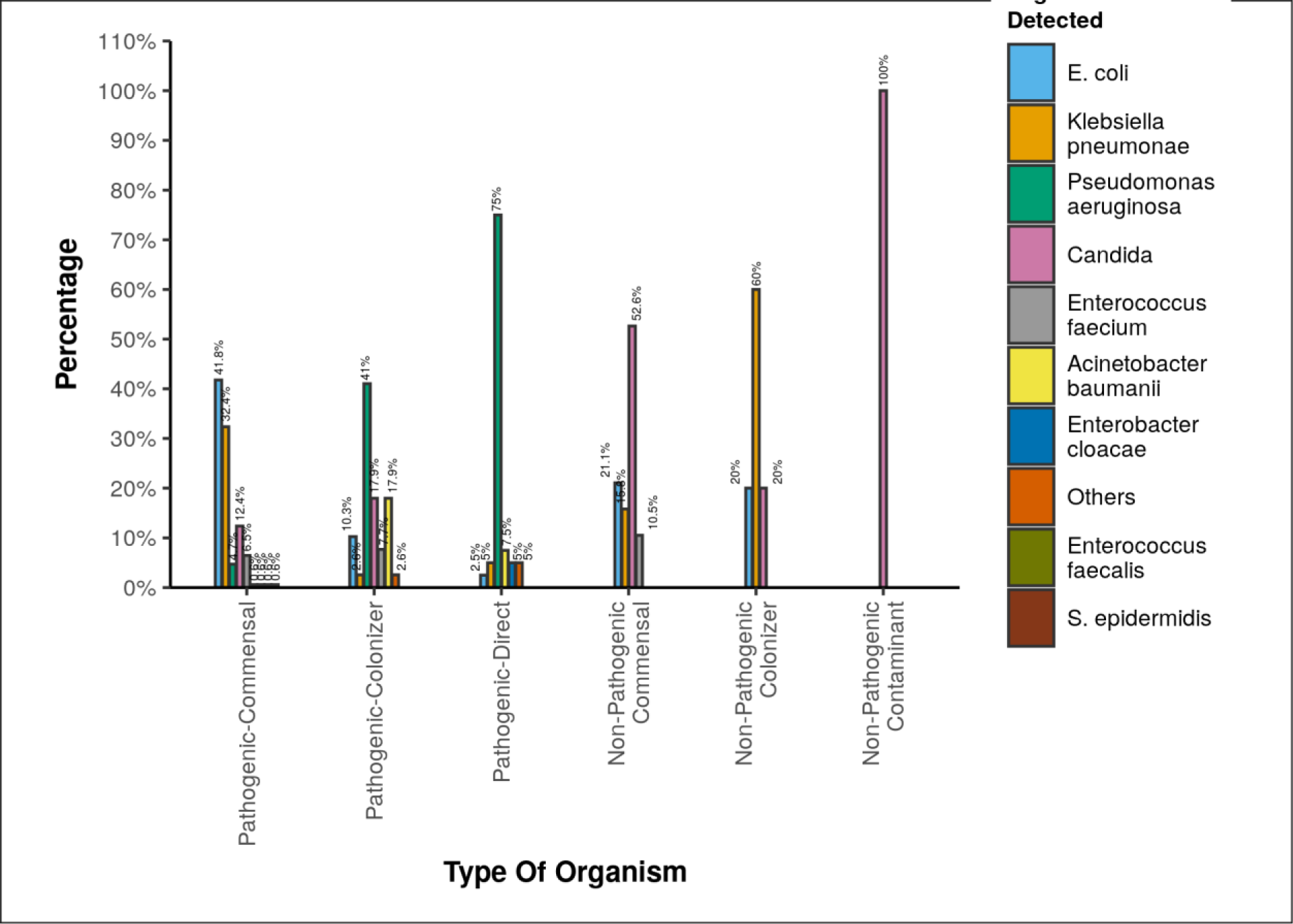
**Association Between Nature of Organism and organism detected**

**Figure 5:**
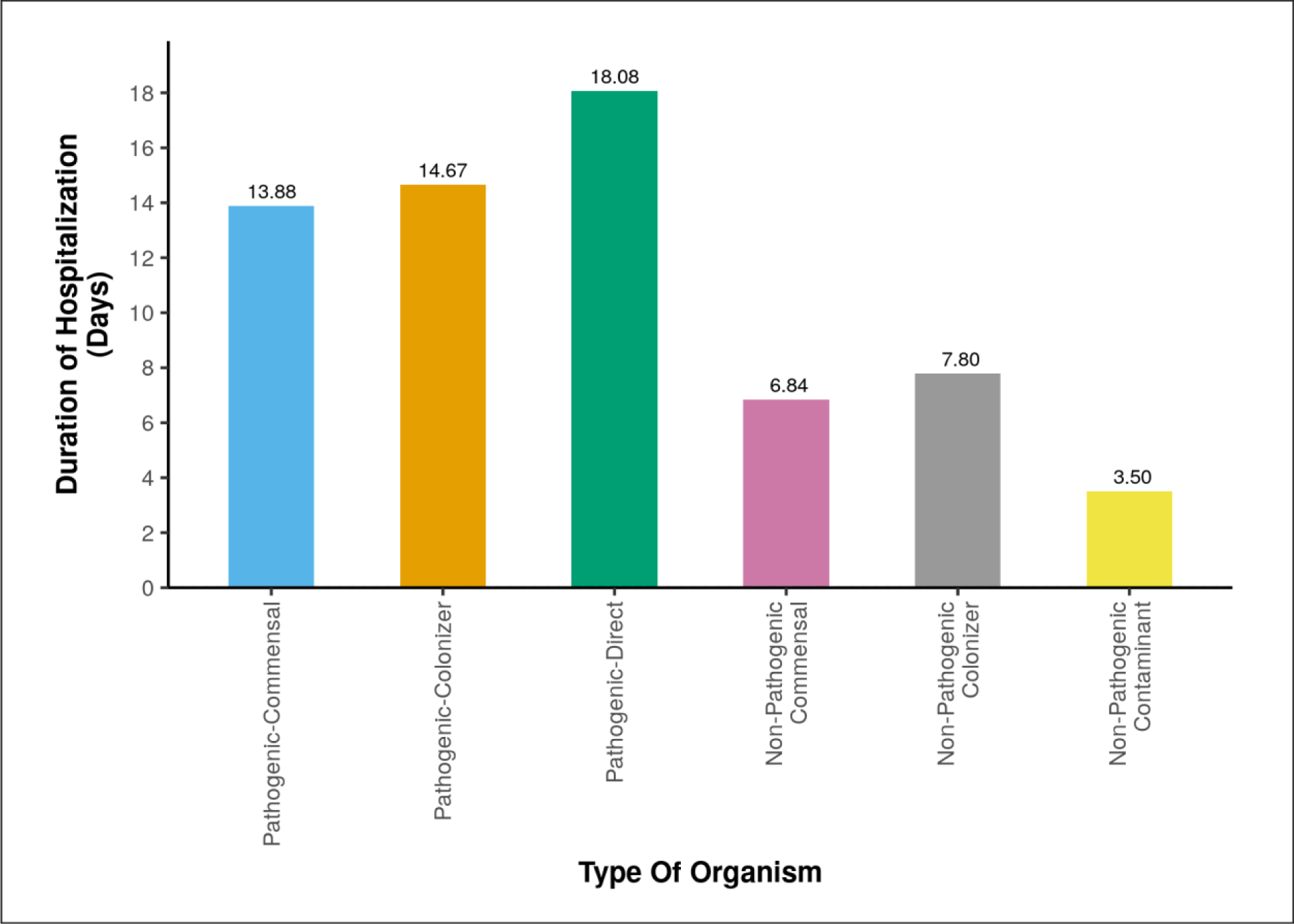
**Association Between Nature of Organism and duration of hospitalization**

**Figure 6:**
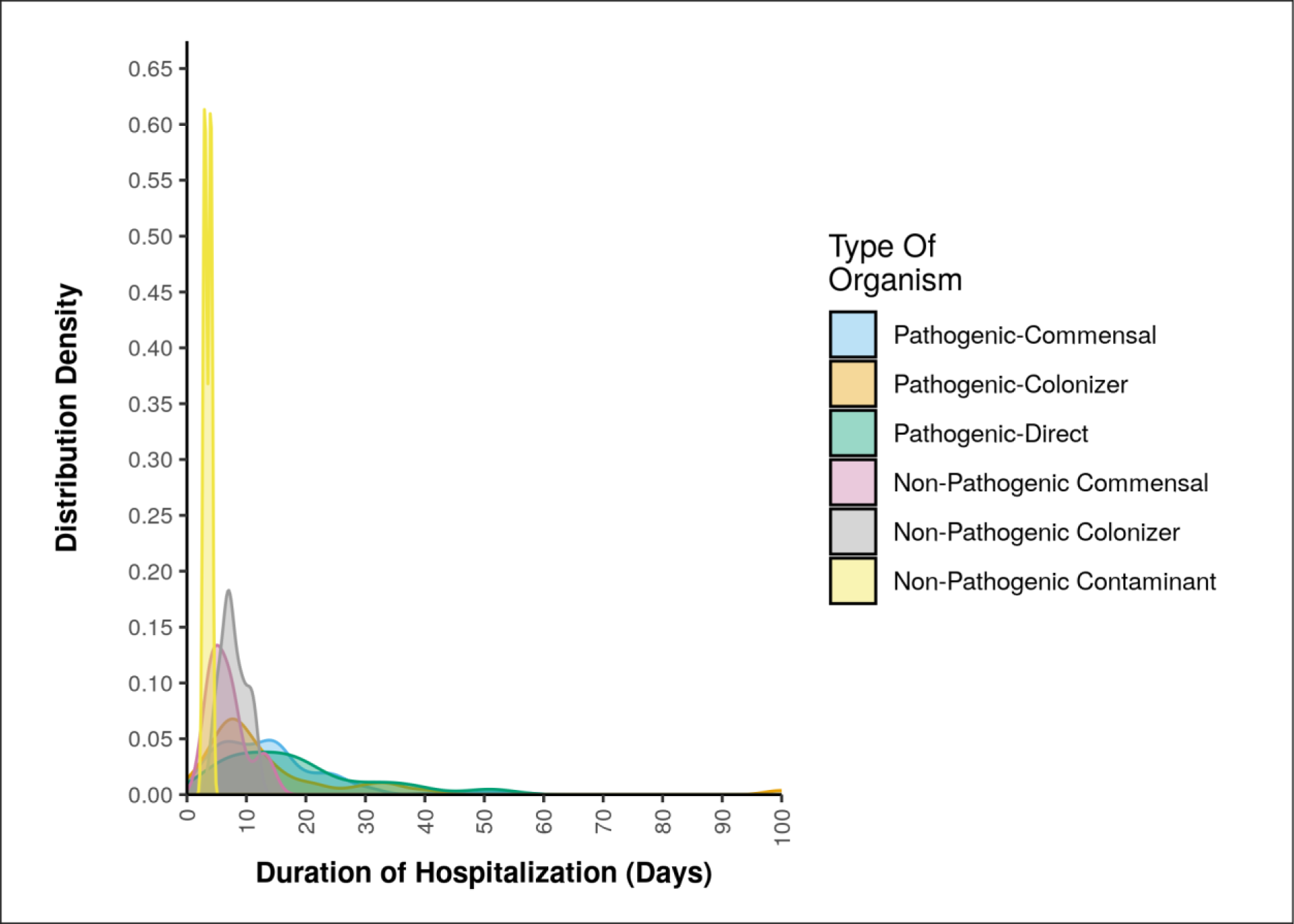
**Association Between Nature of Organism and duration of hospitalization**

**Figure 7:**
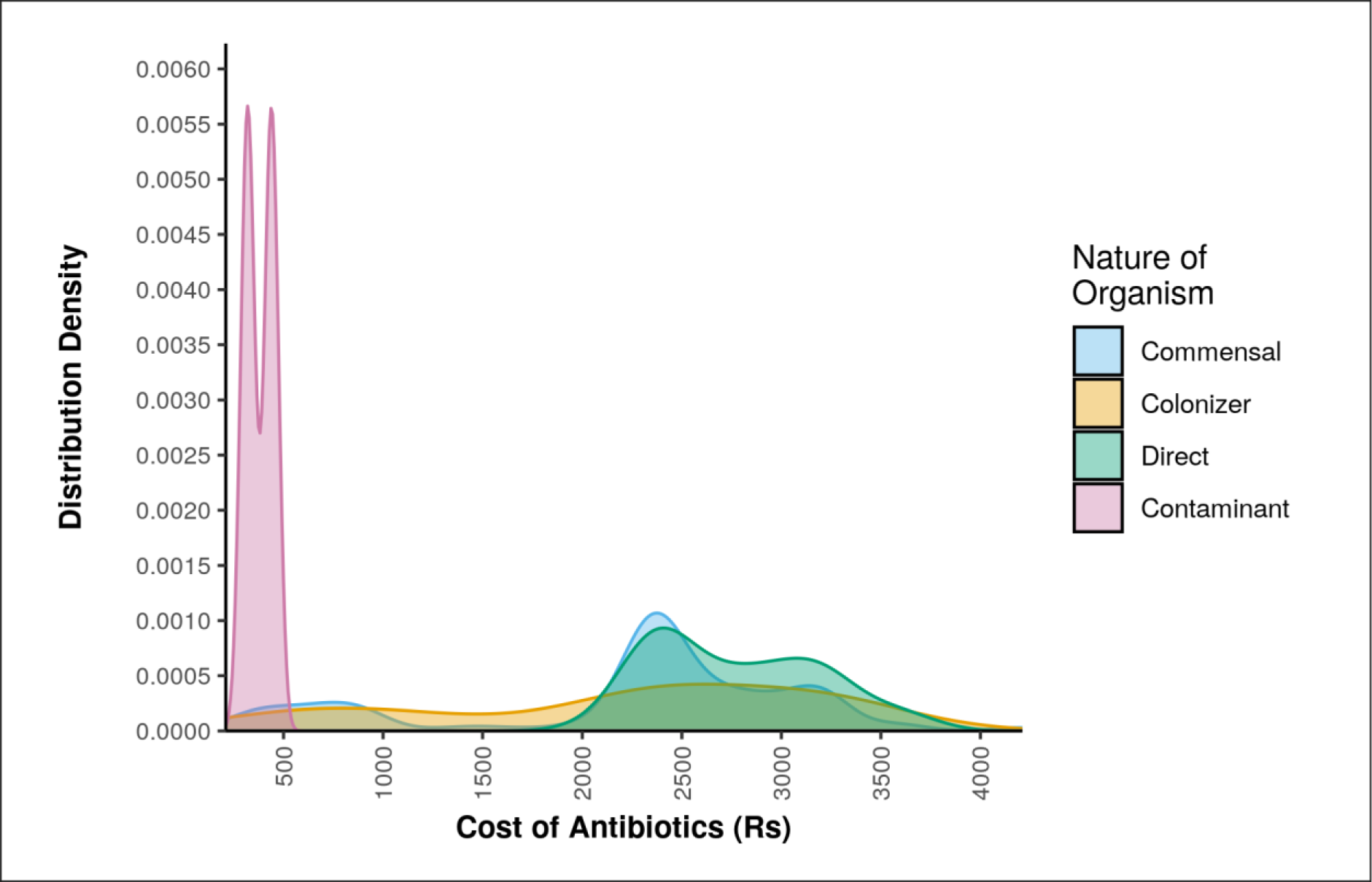
**Association Between Nature of Organism and cost of antibiotics**

### Pathogenic vs. Non-Pathogenic Outcomes

#### Pathogenic Cases (N=249)

- Prevalence:90.54%
- Characteristics

Average Age: 47.32 ± 16.96 years
Gender Distribution: M:F - 156 (62.7%):93 (37.3%)
Average Days of Hospital Stay: 14.68 ± 11.55
Average Days of ABX Usage: 6.81 ± 2.02
Average Expense on ABX: $29.12±9.22
- Mortality: 10 (3.6%)
- Organisms within Pathogenic Cases:

E.coli: 76 cases (30.5%)
K. pneumoniae: 58 cases (23.3%)
Pseudomonas: 54 cases (21.7%)
Other: 24.5%

### Pathogenic Commensal Cases (170

Prevalence: 61.81%
Characteristics

- Average Age: 48.15 ± 16.72
- Gender Distribution: 100:70
- Average Days of Hospital Stay: 13.88 ± 9.65
- Average Days of ABX Usage: 6.62 ± 1.77
- Average Expense on ABX: $28.46±9.33
Mortality: 9 (3.2%)

Pathogenic Colonizer Cases (39):

- Prevalence: 14.18%
- Characteristics

Average Age: 50.36 ± 20.53
Gender Distribution: 28:11
Average Days of Hospital Stay: 14.67 ± 16.95
Average Days of ABX Usage: 7.36 ± 2.65
Average Expense on ABX: $27.69±11.02
- Mortality: 1 (0.36%)

### Direct Pathogenic Cases (40)

- Prevalence: 14.54%
- Characteristics

Average Age: 40.83 ± 12.34
Gender Distribution: 32:8
Average Days of Hospital Stay: 18.08 ± 12.21
Average Days of ABX Usage: 7.05 ± 2.23
Average Expense on ABX: $33.28±5.01

### Non-Pathogenic Cases (N=26)

- Prevalence: 9.45%
- Characteristics

Average Age: 50.88 ± 13.42
Gender Distribution: M:F - 16 (61.5%):10 (38.5%)
Average Days of Hospital Stay: 6.77 ± 3.06
Average Days of ABX Usage: 6.23 ± 1.34
Average Expense on ABX: $11.79±11.02
- Mortality: 0

### Non-Pathogenic Commensal Cases (19)

- Prevalence: 6.9%
- Characteristics

Average Age: 50.21 ± 13.67
Gender Distribution: 11:8
Average Days of Hospital Stay: 6.84 ± 3.22
Average Days of ABX Usage: 6.53 ± 1.31
Average Expense on ABX: $11.59±11.07
- Organisms within Non-Pathogenic Cases:

Candida: 13 cases (50.0%)
E.coli: 5 cases (19.2%)
Klebsiella: 6 cases (23.1%)
Other: 7.7%

### Non-Pathogenic Colonizer Cases (5)

- Prevalence: 1.81%
- Characteristics

Average Age: 55.20 ± 15.75
Gender Distribution: 4:1
Average Days of Hospital Stay: 7.80 ± 2.28
Average Days of ABX Usage: 5.00

## DISCUSSION

### Novelty and Significance of the Study

This study marks a pioneering exploration into the clinical variances of urinary isolates, specifically distinguishing between pathogenic and non-pathogenic organisms, within the unique healthcare landscape of North India. By delving into the nuances of subcategories among both pathogenic and non-pathogenic cases, this research stands as the first of its kind, offering unprecedented insights into the economic and clinical implications of urinary tract infections (UTIs) in a tertiary healthcare setting.

### Comparative Analysis of Pathogenic and Non-Pathogenic Cases

The comprehensive analysis reveals a substantial predominance of pathogenic cases, constituting 90.54% of the cohort. This highlights the paramount clinical significance of UTIs caused by pathogenic organisms. The economic impact, characterized by prolonged hospital stays, increased antimicrobial usage, and higher associated costs, emphasizes the urgency of effective management strategies.

Noteworthy is the identification and analysis of subtypes within pathogenic cases, including Commensal, Colonizer, and Direct cases. Each subtype exhibits distinct demographic patterns, length of hospital stay, antimicrobial usage, and financial implications. This granularity in understanding pathogenic cases contributes significantly to tailored patient care and resource optimization.

Conversely, non-pathogenic cases, though representing a smaller percentage (9.45%), demonstrate variability in hospitalization duration and antimicrobial expenses. The identification of subtypes within non-pathogenic cases, such as Commensal, Colonizer, and Contaminants, underscores the complexity of UTIs even in seemingly milder cases.

### Clinical Implications and Recommendations

The clinical implications of this study extend beyond the apparent severity of UTIs. The findings advocate for a nuanced approach to patient care by recognizing the potential clinical impact of seemingly benign non-pathogenic isolates. Given the rise of antimicrobial resistance and the economic burden associated with unnecessary antibiotic usage, the identification of non-pathogenic cases gains paramount importance.

This study underscores the significance of distinguishing between pathogenic and non-pathogenic cases to guide clinical decision-making. It is imperative to develop strategies that aid in the identification of non-pathogenic cases, allowing for a judicious approach to antimicrobial prescription. Avoiding antibiotics in non-pathogenic cases not only prevents unnecessary patient exposure but also contributes to global efforts in combating antimicrobial resistance and reduces the economic burden on healthcare systems worldwide.

## CONCLUSION

In conclusion, this study provides a ground-breaking understanding of clinical variances in urinary isolates in North India’s tertiary healthcare. By comparing and contrasting pathogenic and non-pathogenic cases, the research illuminates the need for customized management strategies. The identification of non-pathogenic cases and the avoidance of unnecessary antimicrobial usage emerge as pivotal interventions to address both clinical and economic implications. As the world grapples with the challenges of antimicrobial resistance and healthcare expenditure, this study serves as a beacon for optimizing patient care and contributing to the global fight against unnecessary antibiotic usage.

## DATA SHARING

It will be made available to others as required upon requesting the corresponding author.

## Data Availability

All data produced in the present study are available upon reasonable request to the authors
All data produced in the present work are contained in the manuscript

## ACKNOWLEDGMENT

None

